# Clinical and Radiological Evaluations of COVID-19 Patients with Anosmia: Preliminary Report

**DOI:** 10.1101/2020.05.20.20106633

**Authors:** Jerome R. Lechien, Justin Michel, Thomas Radulesco, Carlos M. Chiesa-Estomba, Luigi A. Vaira, Giacomo De Riu, Leigh Sowerby, Claire Hopkins, Sven Saussez

## Abstract

**Objective:** To investigate clinical and radiological features of olfactory clefts of patients with mild coronavirus disease 2019 (COVID-19).

**Methods:** Sixteen COVID-19 patients were recruited. The epidemiological and clinical data were extracted. Nasal complaints were assessed through the sino-nasal outcome test 22 (SNOT-22). Patients underwent psychophysical olfactory testing, olfactory cleft examination and CT-scan.

**Results:** Sixteen anosmic patients were included. The mean Sniffin’Sticks score was 4.6±1.7. The majority of patients had no endoscopical abnormality, with a mean olfactory cleft endoscopy score of 0.6±0.9. The olfactory clefts were opacified in 3 patients on the CT-scan. The mean radiological olfactory cleft score was 0.7±0.8. There were no significant correlations between clinical, radiological and psychophysical olfactory testing.

**Conclusion:** The olfactory cleft of anosmic COVID-19 patients is free regarding endoscopic examination and imaging. The anosmia etiology would be not related to edema of the olfactory cleft.

**Level of Evidence:** 4

## Introduction

Since the onset of the coronavirus disease 2019 (COVID-19) pandemic in Europe, many otolaryngologists have reported a significant increase in patients with a sudden loss of smell.^1,2^ The olfactory dysfunction may be associated with respiratory symptoms and fever, but also in isolation or in paucisymptomatic disease. Nowadays, olfactory dysfunction has been recognized as a key symptom of COVID-19, with more than 66% of patients in Europe and U.S reporting some degree of hyposmia,^1,3^ leading to an inclusion of olfactory dysfunction in the diagnostic criteria by the World Health Organization. The olfactory disorder has been reported to occur before (11.8%), at the same time (22.8%) or after (65.4%) the presentation of other symptoms.^1^

Since this recognition of the high prevalence of loss of sense of smell reported by COVID-19 patients, there has been much speculation regarding the underlying pathophysiological mechanism. Different theories have been proposed ranging from conductive loss due to obstruction of the olfactory cleft^4^ to central mechanisms relating to the known neurotropic properties of the human coronavirus.^5^

Due to restrictions on travel and overwhelming demands placed on healthcare resources, there have been few reports in the literature of clinical and radiological investigations in these patients. We set out to undertake detailed evaluation of a series of patients with COVID-19 anosmia with the aim of further elucidating the underlying cause of the anosmia.

## Methods

The Jules Bordet Institute approved the study protocol (Central Ethics Committee, IJB-0M011-3137). Patients were invited to participate and informed consent was obtained once inclusion criteria were met.

### Setting

Adults with confirmed COVID-19 and self-reported sudden-onset olfactory dysfunction were recruited through a public call from the Department of human Anatomy of the University of Mons (Mons, Belgium). Only mild-to-moderate patients were included. The patients were defined as mild-to-moderate if they did not require hospitalization to manage the infection. The COVID-19 diagnosis was based on the WHO interim guidance and symptoms of disease. The diagnosis was confirmed through nasopharyngeal swab to identify severe acute respiratory coronavirus-2 (SARS-CoV-2) and related reverse transcription polymerase chain reaction (RT-PCR) analysis. In case of negative RT-PCR, serology was performed (Zentech, University of Liege Lab, Liege, Belgium). The details about the diagnosis procedure were reported in a previous publication.^6^ Note that individuals with a history of olfactory dysfunction before the pandemic (e.g. chronic rhinosinusitis, history of nasal surgery, head & neck trauma, or degenerative neurological disease) were carefully excluded.

### Epidemiological & Clinical outcomes

To minimize the risk of exposure for study personnel, the clinical and epidemiological characteristics of patients were electronically collected via an online questionnaire developed with Professional Survey Monkey® (San Mateo, California, USA). Demographic data including gender, age, and patient comorbidities were collected. Symptoms were evaluated through a 4-point scale ranging from 0 (no symptom) to 4 (severe symptoms). The nasal symptoms were evaluated through the French version of the sino-nasal outcome test 22 (SNOT-22).^7^

### Psychophysical Olfactory Evaluation

The psychophysical olfactory evaluations were performed using the identification Sniffin’ Sticks test (Medisense, Groningen, The Netherlands), which is a validated objective test of olfactory dysfunction.^8^ Sixteen scents were presented via a pen device to patients for 3 seconds followed by a forced choice from 4 given options with a total possible score of 16 points. Regarding results, patients were classified as anosmic (score 8 or below), hyposmic (score between 9-11), or normosmic (score between 12-16).

### Computed tomography (CT-scan)

CT-scan images were independently analyzed by 3 raters, applying a Lund-Mackay style scoring system:^9,10^ 0 for completely clear, 1 for partially opacified and 2 for completely opacified. The olfactory cleft radiological evaluations were made through a “semiquantitative” 0-3 Likert scale.^11^ Where there was disagreement between scores, the modal score was used. Each side was scored separately, with a total score ranging from 0 to 4.

### Olfactory Cleft Endoscopy

Patients underwent rigid endoscopy with findings scored using the olfactory cleft endoscopy scale,^11^ which is a validated scale reporting the findings of discharge, polyps, oedema, scarring or crusting on a scale of 0, 1 or 2 on each side, giving a total score ranging from 0 to 40. Endoscopies were performed by JRL and SS using full personal protective equipment.

### Statistical Analyses

Statistical analyses were performed using the Statistical Package for the Social Sciences for Windows (SPSS version 22,0; IBM Corp, Armonk, NY, USA). The relationship between clinical and olfactory outcomes was analyzed through non-parametric test using Spearman correlation for scale data. We investigated all potential associations between nasal obstruction, rhinorrhea, endoscopy and radiology scores and the occurrence of olfactory disorder (Sniffin’ Sticks test). A level of significance of p<0.05 was used.

## Results

A total of 16 patients underwent complete evaluation with regard to endoscopy, CT-scan and objective olfactory testing. The mean age was 36 ± 10.1 years. There were 8 females (50%) and 8 males (Table 1). All were non-smokers. Ten patients (62.5%) did not report nasal obstruction, 5 reported mild (31.3%) and 1 moderate obstruction (6.3%) (Table 2). The mean SNOT-22 score was 28.8 ±18.0, with a mean score of 4.4 ± 0.7 for the item decreased sense of smell/taste (Table 3). All patients self-reported complete loss of sense of smell at presentation, which was confirmed through Sniffin’Sticks tests, with a mean Sniffin’Stick score of 4.6 ± 1.7 (Table 4).

**Table 1:** Patient Characteristics.

footnotes: Abbreviations: N=number; SD=standard deviation.
| Characteristic | N (%) |
| --- | --- |
| <u>Age</u> |  |
| Mean (Mean, SD) - yo | 36.0 ± 10.1 |
| <u>Gender (N - %)</u> |  |
| Female | 8 (50.0) |
| Male | 8 (50.0) |
| <u>Addictions (N - %)</u> |  |
| Non-smoker | 16 (100) |
| Allergic patients | 1 (6.3) |
| <u>Comorbidities</u> |  |
| Hypothyroidism | 1 (6.3) |
| Psoriasis | 1 (6.3) |
| Obstructive apnea syndrome | 1 (6.3) |
| Diabetes | 1 (6.3) |
| Hepatic insufficiency | 1 (6.3) |
| <u>General Symptoms</u> |  |
| Asthenia | 7 (44.) |
| Loss of appetite | 5 (31.3) |
| Headache | 5 (31.3) |
| Arthralgia | 4 (25.0) |
| Myalgia | 4 (25.0) |
| Diarrhea | 4 (25.0) |
| Abdominal pain | 3 (18.8) |
| Cough | 3 (18.8) |
| Fever | 2 (12.5) |
| Chest pain | 2 (12.5) |
| Nausea/vomiting | 2 (12.5) |
| Urticaria | 2 (12.5) |
| Sticky Sputum | 1 (6.3) |
| Conjunctivitis | 1 (6.3) |

**Table 2:** Severity of Otolaryngological Symptoms developed over the Clinical Course of the Disease (Percent of patients).

footnotes: The symptoms severity was assessed with a 4-point scale (from no problem (0) to Severe problem (4)).
| **Symptoms** | No symptom | Mild | Moderate | Severe | Very Severe |
| --- | --- | --- | --- | --- | --- |
| Nasal obstruction | 10 (62.5) | 5 (31.3) | 1 (6.3) | 0 (0) | 0 (0) |
| Rhinorrhea | 14 (87.5) | 1 (6.3) | 1 (6.3) | 0 (0) | 0 (0) |
| Postnasal drip | 12 (75.0) | 2 (12.5) | 1 (6.3) | 0 (0) | 0 (0) |
| Throat pain | 10 (62.5) | 2 (12.5) | 1 (6.3) | 1 (6.3) | 0 (0) |
| Facial pain | 16 (100) | 0 (0) | 0 (0) | 0 (0) | 0 (0) |
| Ear pain | 14 (87.5) | 1 (6.3) | 0 (0) | 1 (6.3) | 0 (0) |
| Dysphagia | 16 (100) | 0 (0) | 0 (0) | 0 (0) | 0 (0) |
| Dyspnea | 16 (100) | 0 (0) | 0 (0) | 0 (0) | 0 (0) |
| Dysphonia | 15 (93.75) | 0 (0) | 0 (0) | 1 (6.3) | 0 (0) |

**Table 3:** Sino-nasal Complaints of Patients with Olfactory Dysfunction.

footnotes: Abbreviations: QOD-NS= short version of Questionnaire of Olfactory Disorders-Negative Statements; SNOT-22= sinonasal outcome test-22.
|  |  |
| --- | --- |
| **SNOT-22** |  |
| Need to blow nose | 1.6 ± 1.1 |
| Nasal blockage | 1.2 ± 1.0 |
| Sneezing | 1.6 ± 1.2 |
| Runny nose | 1.0 ± 1.3 |
| Cough | 1.0 ± 1.2 |
| Post-nasal discharge | 0.2 ± 0.4 |
| Thick nasal discharge | 0.2 ± 0.4 |
| Ear fullness | 0.4 ± 0.6 |
| Dizziness | 0.4 ± 0.6 |
| Ear pain | 0.4 ± 0.7 |
| Facial pain/pressure | 0.6 ± 1.1 |
| Decreased sense of smell/taste | 4.4 ± 0.7 |
| Difficulty falling asleep | 1.2 ± 1.7 |
| Wake up at night | 1.4 ± 1.5 |
| Lack of a good night's sleep | 2.0 ± 1.6 |
| Wake up tired | 1.6 ± 1.6 |
| Fatigue | 1.8 ± 1.5 |
| Reduced productivity | 1.5 ± 1.6 |
| Reduced concentration | 1.4 ± 1.2 |
| Frustrated/restless/irritable | 1.8 ± 1.7 |
| Sad | 1.4 ± 1.7 |
| Embarrassed | 1.3 ± 1.5 |
| **SNOT-22 total score** | **28.8 ± 18.0** |
| **Short version QOD-NS items** |  |
| Changes in my sense of smell isolate me socially. | 1.3 ± 1.0 |
| The problems with my sense of smell have a negative impact on my daily social activities | 1.1 ± 1.1 |
| The problems with my sense of smell make me more irritable | 1.5 ± 1.1 |
| Because of the problems with my sense of smell, I eat out less | 1.1 ± 1.1 |
| Because of the problems with my sense of smell, I eat less than before (loss of appetite) | 1.3 ± 1.0 |
| Because of the problems with my sense of smell, I have to make more effort to relax | 1.6 ± 1.0 |
| I'm afraid I'll never be able to get used to the problems with my sense of smell. | 0.5 ± 0.7 |
| **Short version QOD-NOS total score** | **8.1 ± 4.0** |

**Table 4:** Endoscopy & Psychophysical Test Features of Patients.

footnotes: Abbreviation: SD=standard deviation.
|  |  |
| --- | --- |
| <u>Anosmia Features</u> |  |
| <u>Sniffin Sticks Tests (Mean, SD)</u> | 4.6 ± 1.7 |
| <u>Olfactory Cleft Endoscopic Scale (Mean, SD)</u> |  |
| Discharge | 0.2 ± 0.6 |
| Polyps | 0.0 ± 0.0 |
| Edema | 0.3 ± 0.7 |
| Crusting | 0.1 ± 0.3 |
| Scarring | 0.0 ± 0.0 |
| **Total Olfactory Cleft Endoscopy Scale** | 0.6 ± 0.9 |
| <u>Lund & Mackay score (Mean, SD)</u> | 0.8 ± 0.9 |

The majority of patients had the CT-scan, endoscopy and olfactory testing performed within 48-72 hours of each other. The longest interval between tests was 3 days. There was a variable interval from the onset of loss of smell and testing due to patient recruitment (mean: 19.8 ± 12.8 days). The mean Lund & Mackay score was 0.8 ± 0.9.

The mean radiological olfactory cleft score was 0.7 ± 0.8; 13 patients were rated as having the olfactory clefts completely (7) or partly (6) clear while 3 patients were completely opacified on CT-scan imaging (Figure 1).

**Figure 1:**
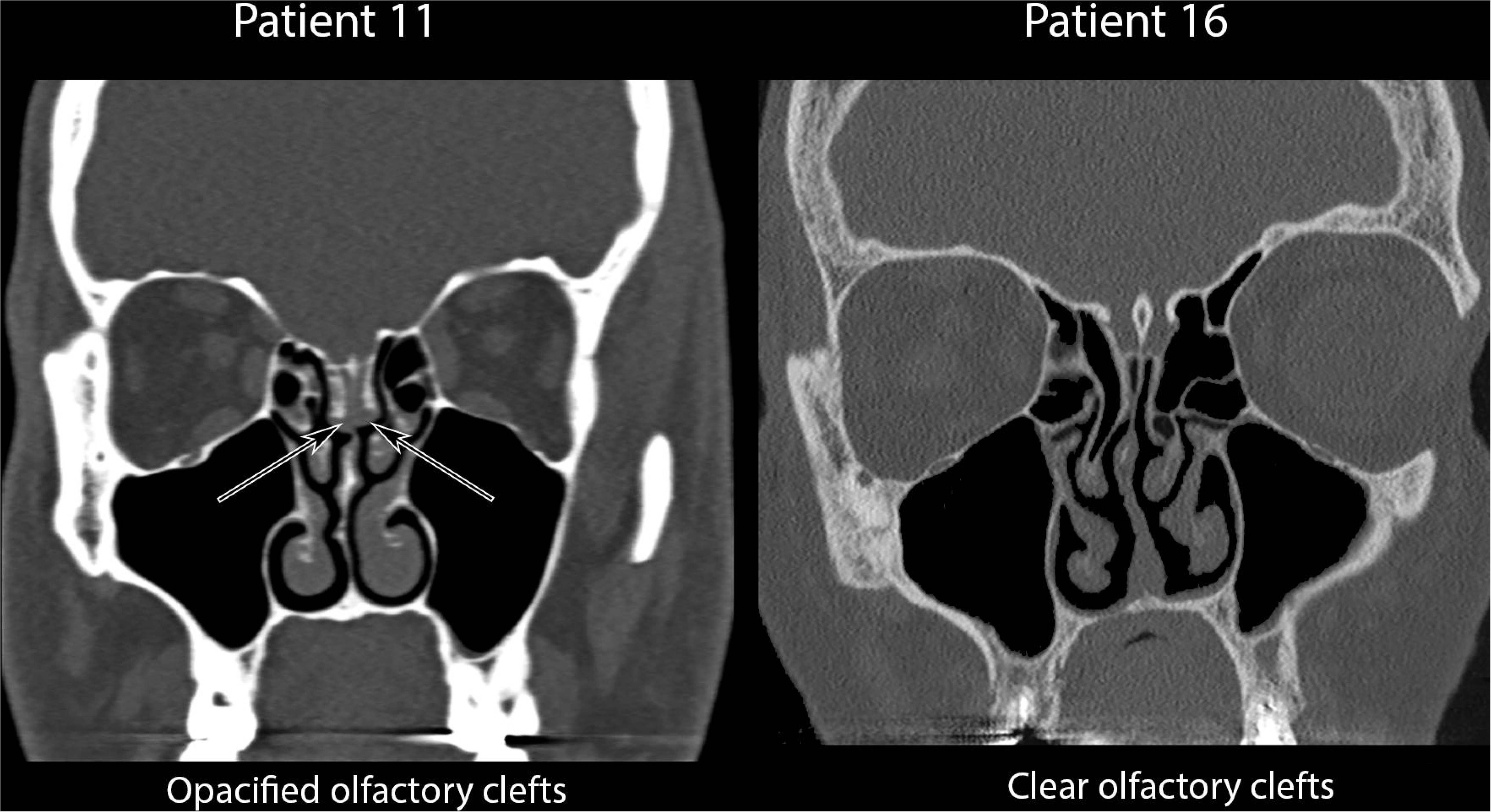
CT-Scan Findings of Patients. **Figure 1 footnotes**: Examples of Patients with opacified olfactory clefts (Patient 11) or patients with clear olfactory clefts (Patient 16).

Endoscopy findings showed that the majority of patients had no visible abnormality, with a mean total olfactory cleft endoscopy score of 0.6 ± 0.9, with edema being the most common finding (mean score of 0.3 ± 0.7) (Table 4). There were no significant correlations between endoscopy, CT-scan findings, SNOT-22, nasal obstruction or Sniffin’Sticks scores. Of all univariate comparisons, only the correlation between CT-scan and Sniffin’Sticks scores approached significance (Rho=-0.46, p=0.09).

## Discussion

To date, there have only been two published cases of COVID-19 anosmia with radiological investigation. The first presented the magnetic resonance imaging (MRI) of a 27 year-old male with sudden onset complete anosmia, but found normal volume and signal intensity of the olfactory bulb with no inflammatory changes in the nasal cavity or olfactory cleft.^12^ We therefore read with interest the case report detailing CT-scan findings in a young female with COVID-19 related anosmia.^4^ The CT-scan demonstrated bilateral opacification of the olfactory clefts; this was proposed as the mechanism of underlying anosmia, with the obstructive inflammation preventing the passage of odorant molecules to the olfactory epithelium. The syndrome of inflammatory obstruction of the olfactory clefts has previously been described by the same team: in their study of 34 patients with complete obstruction of the olfactory clefts, patients showed severe deficits in olfactory function, both in terms of detection and identification.^13^

Indeed, there is further support for their hypothesis in the literature; in patients with chronic rhinosinusitis with nasal polyps, there is a significant correlation between olfactory cleft opacification and results of psychophysical measures of olfactory function.^14,15^ Furthermore, a study using self-reported reduction in sense of smell reported that total, but not partial opacity, of the olfactory cleft correlated with decreased sense of smell.^16^ In a previous experimental model of coronavirus-induced common cold in young adults, impairment in olfactory function was correlated to nasal obstruction.^17^ However in this study, while the subjects developed hyposmia, none became anosmic.^17^

We have previously evaluated a group of 86 patients presenting with initial onset loss of sense of smell, with confirmed COVID-19 infection on either PCR or serology.^6^ On psychophysical olfactory testing, 52% of patients were anosmic. Although 46% of our cohort reported nasal obstruction to some degree, we found no significant correlation between the severity of nasal obstruction and that of the olfactory loss.^18^ However, we note that the patient reported by Eliezer *et al*., also failed to report nasal obstruction, and, therefore, sought to confirm whether our cohort may have localized obstruction of the olfactory cleft.^4^ Due to fear of contamination and ongoing travel restrictions, we have been able to undertake complete evaluation in only 16 patients. While all patients were confirmed to be anosmic on testing, 7 patients had completely or partly clear olfactory clefts on both sides, 6 had partial bilateral opacification and only 3 patients had bilateral complete opacification of the olfactory cleft. Endoscopy did not demonstrate congestion of the olfactory cleft. Thus, the mechanism proposed by Eliezer *et al*. cannot account for the anosmia in the majority of patients, likely supporting a sensorineural etiology. There was a trend towards significance suggesting a weak correlation between olfactory cleft obstruction and olfactory dysfunction, suggesting that, when present, the obstruction may increase the severity of olfactory dysfunction in some patients. How much the observed olfactory opacification contributes to the severity of olfactory dysfunction, and how long it persists deserves further investigation as it may help determine the potential role, if any, for treatments such as topical steroids in the management of olfactory loss. However, Troitier *et al*. report that such patients are usually unresponsive to topical therapies.^13^

The neurotropic potential for human coronavirus has been previously demonstrated. Netland *et al*. demonstrated on transgenic mice expressing the SARS-CoV receptor (human angiotensin converting enzyme 2 – ACE2) that SARS-CoV may enter the brain through the olfactory bulb,^5^ and from there propagate by direct axonal transmission.^19^ Reduced olfactory bulb and olfactory cortex volume has been demonstrated in patients with persistent post viral anosmia.^20^ However, expression of ACE2 has been demonstrated by the sustentacular cells of the olfactory epithelium, although not by the olfactory neurons themselves.^21^ Thus it has been proposed that damage to the supporting epithelium may be responsible for the observed, and often transient olfactory loss. In our small cohort, the majority of patients remain anosmic at their second assessment with psychophysical tests (duration from onset of anosmia 27 – 49 days), suggesting the role of a central mechanism.

The limitation of this study is the small numbers and variable duration from onset of symptoms to CT-scan and endoscopy, unavoidable due to the restrictions on both travel and access to imaging and endoscopy. In order to mitigate against this, Sniffin’Sticks testing was repeated within 10 days of imaging, and on the day of endoscopy. Furthermore, in order to minimize contact time and reduce risk of contamination, only the identification part of the Sniffin’Sticks assessment was made; however as identification was shown to be severely impaired in obstructed olfactory cleft disease,^13^ we believed this would have sufficient sensitivity. Future studies should ideally include MRI, although both reported cases have failed to demonstrate any significant findings in the acute phase, and as changes in the olfactory bulb and cortex are related to the duration of post-viral olfactory loss,^20^ such changes will likely only be detected with delayed MRI.

## Conclusion

This is the first study to report both endoscopic and radiologic imaging in a series of patients with COVID-19 related anosmia. Our findings suggest that while obstruction of the olfactory cleft may play a small role in increasing the severity of the olfactory dysfunction, it does not appear to be the primary underlying mechanism.

## Data Availability

Data available with corresponding author approval

## Acknowledgments

FRMH & UMONS.

## Notes

**Funding**: FRMH & UMONS Grants.

**Conflict of interest statement**: The authors have no conflicts of interest

### Competing Interest Statement

The authors have declared no competing interest.

### Clinical Trial

IJB-0M011-3137

